# Forecasting COVID-19 infection trends and new hospital admissions in England due to SARS-CoV-2 Variant of Concern Omicron

**DOI:** 10.1101/2021.12.29.21268521

**Authors:** Alberto Giovanni Gerli, Stefano Centanni, Joan B Soriano, Julio Ancochea

## Abstract

**Objectives:** On November 26, 2021, WHO designated the variant B.1.1.529 as a new SARS-CoV-2 variant of concern (VoC), named Omicron, originally identified in South Africa. Several mutations in Omicron indicate that it may have an impact on how it spreads, resistance to vaccination, or the severity of illness it causes. We used our previous modelling algorithms to forecast the spread of Omicron in England.

**Design:** We followed EQUATOR’s TRIPOD guidance for multivariable prediction models.

**Setting:** England.

**Participants:** Not applicable.

**Interventions:** Non-interventional, observational study with a predicted forecast of outcomes.

**Main outcome measures:** Trends in daily COVID-19 cases with a 7-day moving average and of new hospital admissions.

**Methods:** Modelling included a third-degree polynomial curve in existing epidemiological trends on the spread of Omicron and a new Gaussian curve to estimate a downward trend after a peak in England.

**Results:** Up to February 15, 2022, we estimated a projection of 250,000 COVID-19 daily cases of Omicron spread in the worse scenario, and 170,000 in the “best” scenario. Omicron might represent a relative increase from the background daily rates of COVID-19 infection in England of mid December 2021 of 1.9 to 2.8-fold. With a 5-day lag-time, daily new hospital admissions would peak at around 5,063 on January 23, 2022 in the worse scenario.

**Conclusion:** This warning of pandemic surge of COVID-19 due to Omicron is calling for further reinforcing in England and elsewhere of universal hygiene interventions (indoor ventilation, social distance, and face masks), and anticipating the need of new total or partial lockdowns in England.

## Text

England has been among the hardest hit countries by COVID-19 worldwide, particularly during the ongoing sixth wave.^1^ There have been several successful attempts to forecast trends of incidence and mortality of COVID-19, most based upon knowledge on viral dynamics from previous pandemics, recent COVID-19 geographical information of diverse granularity, and newly discovered viral characteristics.^2,3^ However, SARS-CoV-2 inherent poor quality RNAm copy-editing gene replication makes it prone to mutate and spontaneously create new variants of concern (VoC),^4^ that adapt to any hostile environment, produce new outbreaks, and modify existing epidemiological projections.^5^

On November 26, 2021, WHO designated the variant B.1.1.529 as a new VoC, named Omicron, originally identified in South Africa, on the advice of WHO’s Technical Advisory Group on Virus Evolution.^6^ This decision was based on the evidence that Omicron has several mutations that may have an impact on how it spreads, resistance to vaccination, or the severity of illness it causes.^7,8^ In particular, in South Africa up to December 2, 2021 it was observed a doubling time for the first 3 days after the wave threshold of ten cases per 100 000 population.^9,10^

In Denmark, considered a European leader in sequencing SARS-CoV-2 VoC, where testing of all positive PCR tests is commonplace, cases of Omicron have been reported to double every second day.^11^ There, almost 75% of those infected by Omicron had received full (two doses of) COVID-19 vaccination already. On the positive side, it appears most Omicron-related COVID-19 cases are mild or even pauci-symptomatic.

We used our previous modelling algorithms,^12,13,14,15^ to forecast the spread of Omicron in England, and report trends in COVID-19 daily cases with a 7-day moving average and of new hospitalizations. We followed EQUATOR’s TRIPOD guidance for multivariable prediction models.^16^ By applying firstly a third-degree polynomial curve in existing epidemiological trends on the spread of Omicron in England, starting from the first 17 days of the Omicron outbreak (from December, 8, 2021), and secondly a Gaussian curve following a parametric growth,^12-15^ we were able to model new infections of COVID-19 in England. Overall, the “best” scenario forecasts up to 170,800 COVID-19 daily infections up to February 15, 2022 while the worse scenario is 257,167 (Figure 1).

**Figure 1:**
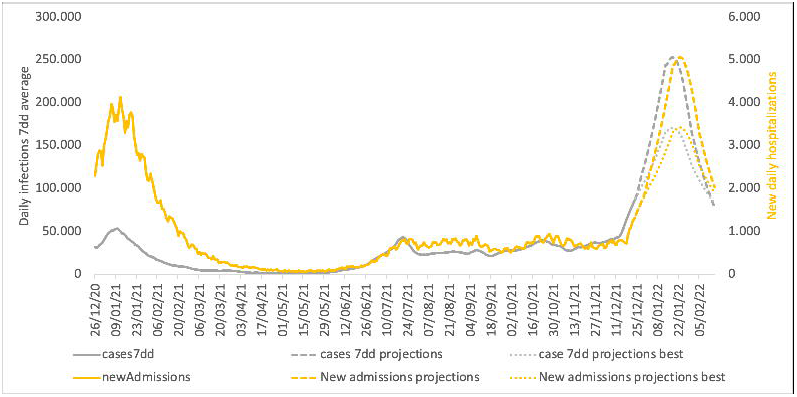
Trends in COVID-19 daily new infections with a seven-day moving average and of new hospital admissions in England, observed and expected up to February 15, 2022

Then we modelled these trends for new COVID-19 hospital admissions using a new Gaussian curve to estimate a downward trend after a peak,^17^ and we obtained the expected curve of new COVID-19 infections in England, and with a 5-day lag time, new hospital admissions. Omicron will likely produce crowding of hospitals in England, as new hospital admissions per day will peak on January 23, 2022, with a range in between 3,416 (“best” scenario) and 5,063 (worse scenario). Both epidemiological indicators will surpass rates observed in the previous five waves in England, unless both individual and group interventions are taking place.

In probability theory, the conditional expectation of any warning system for an eventual surge of an infectious outbreak, as could happen with Omicron substituting other SAR-CoV-2 VoC, modifies (reduces) the eventual magnitude of the event itself.^18^ Given preliminary evidence from South Africa, our forecast anticipates a large COVID-19 burden increase in England despite the high levels of vaccination.^19^ Therefore, this warning is calling for further reinforcing of universal hygiene interventions (indoor ventilation, social distance, and face masks), and anticipating the need of new lockdowns,^11^ the latter being extremely detrimental to the economy.

All viruses change in time and space by natural or artificial Darwin’s selection, and survival of the fittest,^20^ due either to high levels of herd immunity or low vaccination coverage, respectively. The toll associated with VoC Omicron underlines WHO’s COVID-19 message that: “No one will be safe, until the entire World is safe (ergo vaccinated)”.

## Data Availability

All data produced in the present study are available upon reasonable request to the authors

## Notes

### Competing Interest Statement

The authors have declared no competing interest.

### Funding Statement

This study did not receive any funding

### Author Declarations

S Africa and England publicly available data

